# Video feedback for young babies and maternal perinatal mental illness: Adaptation, feasibility and qualitative interviews

**DOI:** 10.1101/2023.09.28.23296278

**Authors:** Kirsten Barnicot, Eloise Stevens, Fiona Robinson, Sarah Labovitch, Rajinder Ballman, Maddalena Miele, Tara Lawn, Sushma Sundaresh, Jane Iles

## Abstract

**Aims/ Background:** We aimed to adapt, test and explore experiences of the video feedback intervention for positive parenting (VIPP) for 2 to 6 month old babies and for mothers experiencing moderate to severe perinatal mental health difficulties.

**Design/ Methods:** The VIPP intervention was adapted to include developmentally appropriate activities and developmental psychoeducation for 2 to 6 month olds, alongside psychoeducation on emotion regulation. Subsequently, the adapted intervention was trialled in 14 mothers experiencing moderate to severe perinatal mental health difficulties (registration <u>ISRCTN64237883</u>). Observational and self-reported pre-post outcome data was collected, and post-intervention qualitative interviews were conducted with participating mothers and clinicians.

**Results:** Consent (67%), intervention completion (79%) and follow-up rates (93%) were high. Effect sizes on pre-post outcome measures indicated large improvements in parenting confidence and perceptions of the parent-infant relationship, and a medium-size improvement in maternal sensitivity. Qualitative interviews suggested that clinicians and mothers were able to use the video feedback to identify young babies’ subtle behavioural cues and moments of mother-infant connection, enhancing maternal sensitivity. Mothers’ initial anxieties about being filmed were overcome by the experience of receiving positive and strengths-focussed feedback, boosting their confidence in themselves as parents. The interviews also generated recommendations for minor modifications to optimise intervention feasibility and acceptability, such as streamlining the information provided on maternal emotion regulation, and allowing increased use of clinical judgement to tailor intervention delivery.

**Conclusion:** VIPP can potentially be beneficial for enhancing maternal sensitivity with very young babies in mothers experiencing perinatal mental health difficulties.

## Introduction

Up to 5% of mothers will experience moderate to severe perinatal mental health problems during pregnancy or postnatally, including bipolar disorder, personality disorders, psychosis, severe anxiety or depressive disorders (Howard et al. 2014; Jones, Chandra, Dazzan & Howard, 2014, National Health Service 2019). These involve significant risk of relapse, suicide, mother-infant relationship difficulties and/or difficulties in day to day functioning and infant care, and can be further complicated by social adversity, child protection concerns, a history of complex trauma and/or intimate partner violence (Howard & Khalifeh 2020, Howard et al. 2014).

Maternal perinatal mental illness increases the risk of future child mental health and behaviour problems, due partly to increased early difficulties in parent-infant relationships (Stein et al. 2014). These may include mothers’ negative perceptions of their relationship with their child, and difficulty responding sensitively to their infant’s communication, which have been linked to subsequent problems in children’s socioemotional development (Aktar et al. 2019; Harries, Smith, Gregg, & Wittkowski 2023; Howard & Khalifeh 2020).

Meta-analysis has shown that the most effective interventions for improving parental sensitivity incorporate direct feedback to the parent, through recording and playing back videos of parent-infant interaction (Bakermans-Kranenburg, van Ijzendoorn, Juffer et al. 2003). Trials in other at-risk groups - mothers with bulimia nervosa, insecure attachment or low sensitivity, and infants with behavioural problems – have shown that the video feedback intervention for positive parenting (VIPP), a six-session parenting programme based on attachment theory (Juffer, Bakermans-Kranenburg, & Van IJzendoorn 2015), is effective in improving parental sensitivity and child mental health (Juffer, Bakermans-Kranenburg, & Van IJzendoorn 2017, O’Farrelly et al. 2021). VIPP involves clinicians analysing videos of parent-child interaction, and sharing feedback with parents on their child’s attachment and exploratory behavioural cues, communication and responses to the world around them.

However, there is just one prior study evaluating its feasibility and acceptability for mothers experiencing moderate to severe or complex perinatal mental health difficulties. Whilst our previous adaptation and trial of the VIPP intervention for mothers with perinatal “personality disorder” (VIPP-PMH) demonstrated feasibility, acceptability, and potential large positive effects on parental sensitivity (Barnicot et al. 2022, Barnicot et al. 2023), the trial highlighted that two further adaptations could be beneficial to increase the utility of the intervention for women with moderate to severe perinatal mental health problems: 1) adapting the intervention for younger babies, and 2) adding additional material to promote mothers’ skills in regulating their emotions in the context of parent-infant interaction. VIPP was originally developed for children aged 6 months and above. However, neurobiological and health economic evidence suggests that earlier intervention can accelerate a positive trajectory in child development (Heckman & Masterov 2007, Shonkoff & Phillips 2000). Being able to offer VIPP-PMH to younger babies would increase its availability for women using community perinatal mental health services, who are frequently referred during pregnancy or soon after giving birth (Barnicot et al. 2023). However, there is no evidence base for the use of VIPP with younger babies, and the standard VIPP intervention contains elements that may be unsuitable for very young babies, such as parents building towers of blocks with their children. Further, babies’ range of behavioural cues is more limited in the first months of life. It is not known whether clinicians will be able to identify a sufficiently rich range of infant cues to feed back to mothers, nor whether mothers will find such feedback helpful at this early stage of their child’s development. Additionally, emotional dysregulation is a transdiagnostic feature of mental health problems (Sloan et al. 2017), associated with parent-infant relationship problems, poor parental sensitivity, child behaviour problems and adolescent emotional dysregulation (Buckholdt, Parra, & Jobe-Shields 2014; Carreras, Carter, Heberle, Forbes, & Gray 2019). Regulating one’s emotions during stressful parent-infant interactions can sometimes be difficult for any parent, and particularly for parents who struggle with emotion dysregulation (Reijman et al. 2016). The utility of VIPP-PMH for the perinatal mental health population may therefore be optimized by adding additional material to support mothers with this challenge.

The present research therefore aimed to:

1. Explore feasibility, acceptability and mother and clinician experiences of an adapted VIPP-PMH intervention for mothers experiencing moderate to severe perinatal mental health difficulties and their 2 to 6 month old infants, and for perinatal mental health clinicians delivering the intervention
2. Establish pre-post effect sizes for maternal sensitivity, parenting stress, parental self-confidence, the parent-child bond, and emotional dysregulation.

## Methods

### Trial Registration

ISRCTN 64237883 https://doi.org/10.1186/ISRCTN64237883

### Ethics Approval

Ethical approval was obtained from the Health Research Authority and the East of England - Cambridge South Research Ethics Committee on 29^th^ June 2021 (ref 21/EE/0139).

### Procedure

Inclusion and exclusion criteria can be found in Table 1. We aimed to recruit 10 to 15 mothers, following typical case series design (Abu-Zidan, Abbas & Hefny 2012). Participants were recruited from three community perinatal mental health services in London, United Kingdom. Eligible mothers were approached by the clinical team, given brief verbal information about the study, and asked for verbal consent to pass their contact details on to the study researcher. The researcher met with interested mothers to obtain written informed consent to participate, and administered the baseline measures. Subsequently, the mothers took part in VIPP-PMH sessions, followed by a final research visit at 5 months post-baseline including a qualitative feedback interview.

**Table 1.** Inclusion and exclusion criteria.

| Inclusion criteria | Exclusion criteria |
| --- | --- |
| 1. Women/ birthing persons | 1. A sibling or co-parent is participating in the trial |
| 2. Experiencing moderate to severe and/or complex perinatal mental health difficulties (as indexed by being under the care of specialist community perinatal mental health services) | 2. The eligible child has a clinical diagnosis of a learning difficulty, developmental disorder or sensory impairment |
| 3. A primary caregiver of a child aged 2 to 3 | 3. The eligible parent has English language or |

Table 1. Inclusion and exclusion criteria
|  |  |
| --- | --- |
| months old at intervention initiation | learning difficulties that are sufficiently severe to prevent them completing study measures even with assistance |
| 4. Age 16 to 65 years old |  |
| 4. Capable of giving informed consent |  |

### Intervention

The intervention comprised six 90-minute sessions using the Video-feedback Intervention to promote Positive Parenting adapted for Perinatal Mental Health (VIPP-PMH). Sessions were delivered every two weeks, primarily in participant homes, with clinic-based or video-conferencing virtual delivery available if desired. In each session, clinicians videoed the parent and child engaging in play and in everyday activities such as feeding, with developmentally appropriate toys provided by the study team. The clinician watched the videos back with the parent in the subsequent session and gave feedback on the child’s interactive and play behaviour from the child’s perspective, highlighting the child’s attachment and exploratory behaviours and reinforcing parents’ sensitive responses. When viewing moments where a parent responded less sensitively or missed the child’s cue, the clinician gave tips for alternative ways of responding and flagged moments of more optimal parental responsiveness elsewhere in the interaction.

In our earlier study, we adapted VIPP for a perinatal mental health population by adding psychoeducation on difficult thoughts and feelings arising from being videoed and receiving feedback, an emphasis on the non-judgemental and child-focussed nature of the intervention, and an opportunity to debrief about any difficult thoughts and feelings at the end of each session. In the present study, we developed the VIPP-PMH intervention further by including age-appropriate activities for babies aged 2 to 6 months, based on activities used when VIPP is delivered with 6 to 12 month olds (VIPP Training and Research Centre, 2015), some earlier preliminary work conducted by the intervention developers to adapt VIPP for young babies (Bakermans-Kranenburg pers. comm.), existing resources on recommended activities for this age range (e.g. NCT 2021, Stoppard 2014), and a survey of 42 mothers about the activities they most and least enjoyed doing with their 2 to 6 month old babies. We included both activities which were likely to be enjoyable, and activities which were likely to be more challenging, since mothers in our previous trial felt it beneficial to receive video feedback on these scenarios (Barnicot et al. 2023). Further, based on a review of existing resources (e.g. Nemours Childrens Health 2021, What to Expect 2021, Zero to Three 2016), we developed additional developmental psychoeducation to be shared with parents at each session on infant communication and play, along the following themes: non-verbal communication (session 1), crying as communication (session 2), mirroring (session 3) sensory play and physical touch (session 4), with sessions 5 and 6 allowing for repetition of previous themes as needed. Finally, psychoeducation on emotion regulation strategies for parent-infant interaction was developed based on existing resources on coping with crying/ early parenthood (e.g. Cry-sis 2021, NSPCC 2021), emotion regulation strategies recommended in dialectical behaviour therapy and mentalization based therapy (Linehan 1993, Bateman & Fonagy 2006), and the results of the adaptation study survey.

Fourteen clinicians from the three participating perinatal mental health teams, including five clinical psychologists, six nursery nurses, two occupational therapists, and a mental health nurse, attended a seven day accredited online training in VIPP with sensitive discipline (the standard intervention for older children) and the adaptations for VIPP-PMH, and completed supervised practice sessions with a non-clinical family, followed by a VIPP-PMH case. Clinicians received supervision from an accredited VIPP supervisor prior to delivery of each intervention session.

### Baseline and Follow-up Measures

The self-report measures administered at baseline and follow-up are described in Table 3 below. Additionally, maternal sensitivity was rated by a trained researcher using the Parent Infant Interaction Observation Scale at both timepoints (Svanberg, Barlow & Tigbe 2013), based on 3 minute clips of mothers interacting with their baby, with the instruction to “be with your baby however you normally would. You can do anything you like.”

### Qualitative Interviews

At the 5-month follow-up, mothers were asked to take part in a qualitative interview about their experiences of the intervention and of the study. The interview schedule asked about parents’ expectations and experiences of the intervention, its appropriateness for their child’s stage of development, what they found helpful or less helpful, and any suggestions for improvement. Clinicians delivering the intervention were also interviewed about their experiences and opinions on feasibility and acceptability, after intervention delivery. The interviews were audio recorded for subsequent transcription.

### Analysis

Pre-post effect sizes were calculated as Hedge’s g coefficients, using the average of standard deviations and the pre-post correlation coefficient (Uanhoro 2021). Qualitative interviews were analysed using reflexive thematic analysis (Braun & Clarke 2019), by Authors 3 and 4, and reviewed by Author 1. Following line-by-line coding of all data in NVivo software (Version 12, QSR, 2018), themes and sub-themes were inductively created to capture the in-depth data on experiences of VIPP-PMH.

### Reflexivity

Through reflexive dialogue throughout the research (Olmos-Vega et al., 2022), we identified that each analyst brought a different perspective to the analysis, with Author 1’s interpretations of the data framed by her previous experiences of researching and delivering VIPP; Author 3 by her relationship with participants formed through research visits; and Author 4 by her own positive and negative experiences of receiving perinatal mental health services.

## Results

### Participants

Of 21 eligible women approached by clinicians, 14 agreed to participate (consent rate 67%), of whom 11 completed all 6 VIPP-PMH sessions (completion rate 79%). One discontinued as she stopped responding to researcher and clinician contact; another two women were unable to complete the intervention due to clinician ill health. Thirteen mothers completed the month-5 follow-up and took part in a qualitative feedback interview (follow-up rate 93%). Twelve clinicians were also interviewed (two could not be reached due to ill health). Participant characteristics are shown in Table 2. The sample was generally highly educated and ethnically diverse.

**Table 2.** Participant characteristics.

|  |  |
| --- | --- |
| Age (years) | 34 (4.3) |
| Ethnicity |  |
| Arabic | 2 (14%) |
| Black | 2 (14%) |
| Hispanic | 2 (14%) |
| South Asian | 1 (7%) |
| White | 7 (50%) |
| Highest education level |  |
| A-levels | 1 (7%) |
| Undergraduate degree | 5 (36%) |
| Postgraduate degree | 8 (57%) |
| Relationship status |  |
| Married | 7 (50%) |
| In an unmarried relationship | 7 (50%) |
| Primary psychiatric diagnosis |  |
| Adjustment disorder | 1 |
| Anxiety and/ or depression | 5 |
| Bipolar affective disorder | 2 |
| Childhood onset emotional disorder | 1 |
| Emotionally unstable personality disorder | 3 |
| Post-traumatic stress disorder | 2 |
| Average time since first contact with psychiatric services (years) M(sd) | 9.0 (6.6) |
| Previous psychiatric emergency department visit | 5 (36%) |
| Previous self-harm | 8 (57%) |
| Scoring above threshold on the McLean BPD Screen | 5 (36%) |

### Pre-post Effect Sizes

Mothers’ average scores on all measures improved between pre- and post-intervention, as shown in Table 3.

**Table 3.**
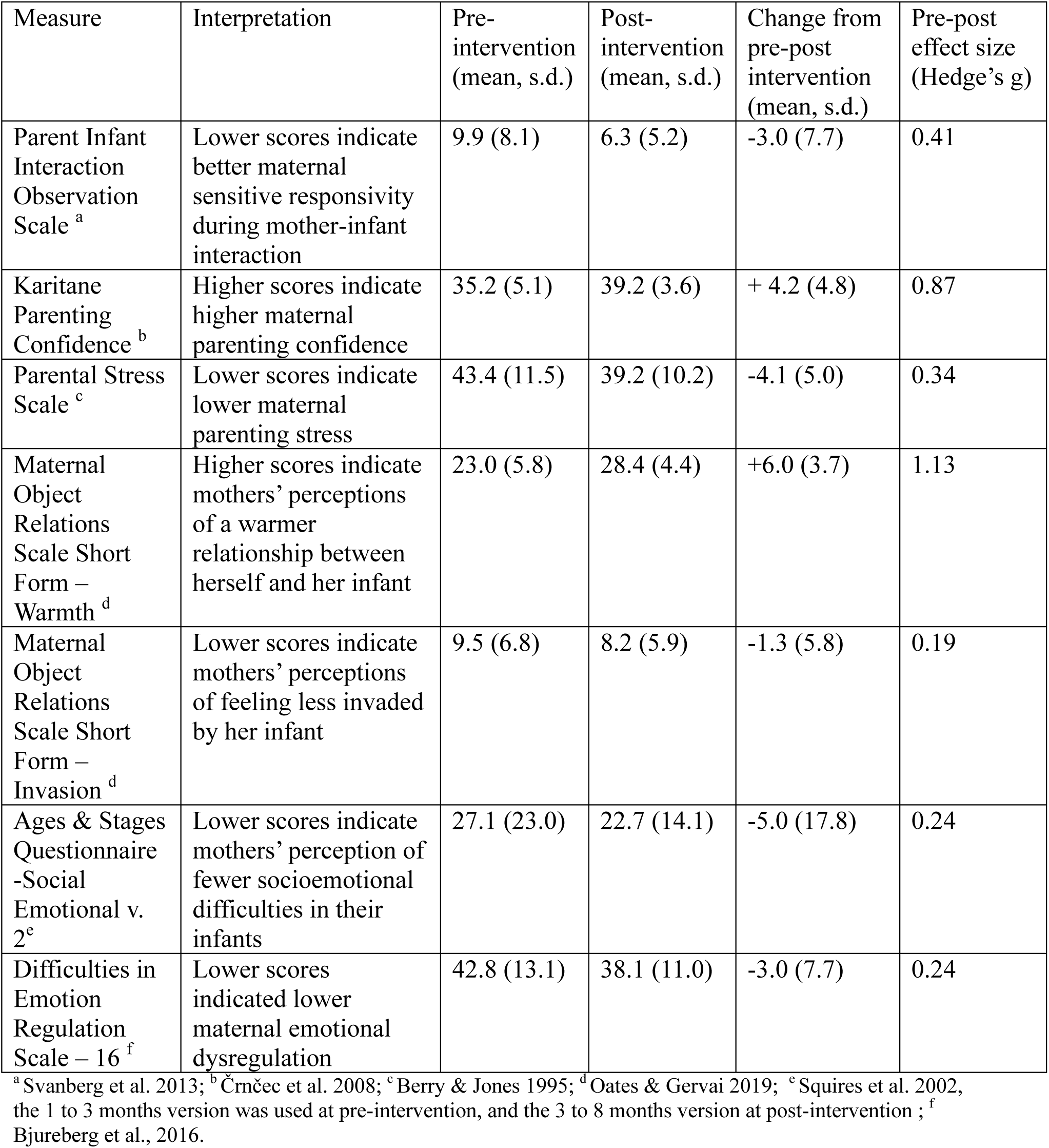
Pre-post outcome measures.

### Experiences of the Intervention

Thematic analysis of the interviews with the mothers and clinicians identified two themes, with a number of sub-themes, focusing on feasibility and acceptability of VIPP-PMH with young babies and with mothers experiencing moderate to severe perinatal mental health difficulties (Figure 1).

**Figure 1.**
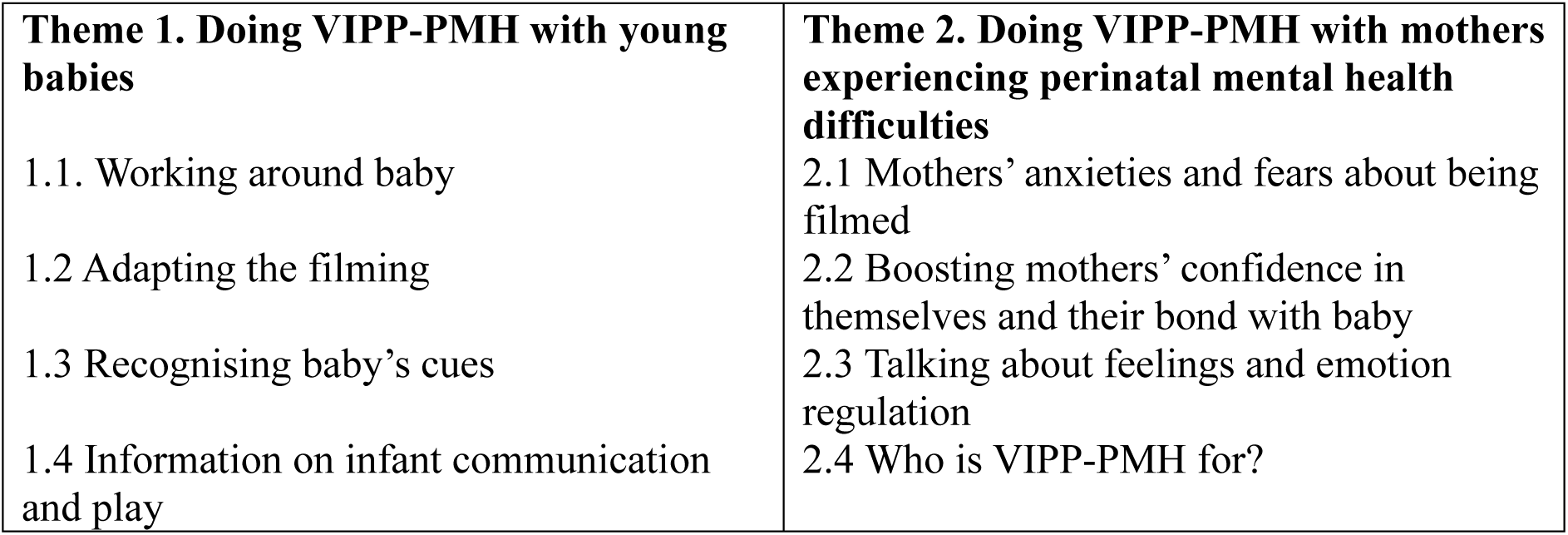
Themes and sub-themes from qualitative feedback interviews with mothers and clinicians.

#### Theme 1. Doing VIPP-PMH with young babies

##### Sub-theme 1.1 Working around baby

Seven clinicians and five mothers explained how flexible and helpful the clinicians had been in trying to fit the sessions around the baby’s timings, and rearranging sessions as needed. This had helped them work around the additional challenges posed by young babies’ unpredictable sleeping and feeding routines, as well as the increased likelihood of their becoming unsettled during the session.

*“I think in the earlier-on sessions ‘cause she was a lot younger, I think we got interrupted a few times because she was hungry …. and then we weren’t able to complete the video ‘cause she fell asleep.”* [P107].

*“A couple of times we had to just change the timing a little bit because she was having a long nap….[Clinician]’d say ‘No it’s ok to give it half an hour and then you can wake her up’. Then I’d text [Clinician] again and say ‘OK I’m ready now.’”* [P111].

##### Sub-theme 1.2 Adapting the filming

Two mothers and one clinician agreed that, for young babies, short filming clips were appropriate due to baby’s more limited interactional repertoire and the increased likelihood of them becoming upset. Further, five clinicians mentioned that filming was easier with younger babies since they were less mobile. The filmed tasks were generally agreed to be developmentally appropriate and mothers mentioned gaining new ideas from them about ways to play with their baby.

*“It gave me ideas as well of things, like I didn’t know she would be at that age yet … so like, we played with bubbles, you know…..it meant that I could go away and do that with her.”* [P104].

However, two clinicians mentioned that a task including a toy for the infant to grasp had been too difficult for the younger babies, but they had nonetheless been able to give the mother some useful feedback about baby’s cues and communication in that context.

*“Baby at that stage wasn’t really confident and skilled in holding and grasping….we just sort of explained, you know…. that you’re looking for those signals from your baby when she’s inviting you to respond and get involved.”* [Clinician of P102].

##### Sub-theme 1.3 Developing a more practiced eye: clinicians and mothers developing skills in recognising baby’s cues

Six clinicians mentioned that, because young babies’ cues are more subtle and their behaviour is more repetitive, it could be challenging to pick out varied moments in the interaction to feed back on. This was felt to particularly be the case when filming feeding, as this was quite a repetitive interactional sequence. However, with experience and with support from their VIPP supervisor, they became more adept at spotting babies’ subtle cues and finding a variety of moments and behaviours in the interaction to comment on.

*“Especially at the beginning, when we’re starting with a two-month-old the communication is very subtle sometimes, so you’re looking just for their eyes, if they’re looking, slight movements of the body…. sometimes I’d watch back and think ‘Ok, I’m not sure what I’m going to comment on here’, and then I’d watch it again and then maybe find a few things, and then when I’d go to supervision we’d find lots of things…. I think it kind of takes a more practiced eye sometimes to see.”* [Clinician of P112].

Eleven mothers and nine clinicians expressed how the video feedback had helped mothers pick up on their baby’s cues and understand more about how and what their baby was communicating to them. For mothers, the most powerful aspect was to see the moments that their baby was responding to them and trying to connect with them, often in subtle ways that they had not previously noticed. This helped mothers to see how important they were to their baby and to see the bond between them.

*“I learnt how to read those slightly smaller, more subtle signals that she gives….like if she was playing with something and then she quickly looked at me and made eye contact with me, something that I maybe wouldn’t necessarily picked up on before.”* [P102]

*“Very simple things like mirroring the mum was so nice. Like the mum loved seeing that, ‘cause she wouldn’t really recognise it when she’s just playing with her baby … his eyes would open when she would talk, if she would sing he would start to move his lips.”*[Clinician of P105].

Five mothers and five clinicians spoke about mothers learning to step back in the interaction and let their baby direct it, slowing down the pace to give the baby more time to process the world around them.

*“I think the biggest thing that I learnt from it was probably giving [Baby] the space to explore things and not feel like I have to intervene all the time….. giving her the space and the time to explore and learn on her own is important.”* [P102].

Both mothers and clinicians remarked that initiating the intervention at two to three months old worked well, as it allowed mothers to recover from childbirth and begin to establish a routine, whilst also enabling mothers to observe a huge development in their children by the time the intervention finished at around six months old.

##### Sub-theme 1.4 Information on infant communication and play

Clinician, but not mothers, were asked in the interviews what they had thought about the additional information on infant communication and play developed for the VIPP-PMH. Eight clinicians felt that this information had been helpful for mothers as it helped them to think about the ways young babies communicate and to see crying as a form of communication, as well as helping them to think about sensory play.

*“I found it was quite a nice thing to add in there. Especially for a parent that, you know, struggling with play or communicating. And even crying, ‘cause a lot of our mums really do struggle with crying. Just to explain, you know, that is really is their main way of communicating.”* [Clinician of P111].

However, one clinician felt that the mother they were working with was already well-versed in this information and did not need to receive it.

#### Theme 2. Doing VIPP-PMH with mothers experiencing perinatal mental health difficulties

##### Sub-theme 2.1 Mothers’ anxieties and fears about being filmed

Twelve mothers and nine clinicians expressed that, exacerbated by their perinatal mental health difficulties, mothers experienced various anxieties about being filmed interacting with their babies. Anxieties ranged from being self-conscious about their appearance, to worrying that the clinician would be judging their parenting. Four mothers also mentioned experiencing self-critical thoughts when watching themselves back on video. However, six mothers and four clinicians explained that mothers felt more comfortable over time as they realised their fears were unfounded.

*“At first, very nerve-wracking, very self-conscious. I had a negative narrative going on for the whole time, like ….’I don’t look a natural mum, we must look like we don’t have a great relationship’….. but then I watched it back and I was like ‘Oh you look fine. We look great, [baby’s] the star.”* [P101]

Seven mothers mentioned that they struggled to be themselves and interact naturally with their baby on camera, as they were very aware of being observed. A few also mentioned feeling like they had to stick the activities suggested by the clinician, and consequently acting differently or being less responsive to their baby than they otherwise would be. Mothers mentioned this getting easier over time and with reassurance from the clinician.

*“I felt like I needed to perform while we’re taking the videos, and she was good at catching that. She kept telling me, ‘You don’t have to do anything, you just need to be 100% yourself, because whatever you’re doing, it’s whatever your baby needs.’”* [P110].

##### Sub-theme 2.2 Boosting mothers’ confidence in themselves and their bond with baby

All 13 interviewed mothers described the clinicians delivering VIPP as kind, caring, non-judgemental, reassuring and making them feel comfortable, which helped to ameliorate their anxieties about being filmed. The focus of the feedback on the baby, and on highlighting positive features of the interaction, also greatly reduced their anxiety.

*“She was just so reassuring. Just really lovely and she would always draw me back to ‘Look how happy [Baby] is. Look at what a happy little baby she is, look at the interaction’… She was just so positive…made me feel very at ease.”* [P111].

Mothers and clinicians described this positive feedback as boosting mothers’ confidence in themselves as parents and helping them to overcome their negative feelings and doubts:

“*I’m frustrated inside because at the time, I’m talking to her and I feel like I’m not really getting there and I’m not really understanding her, but then when I was watching the videos… actually, I was doing great! Actually she did this particular hand movement and I knew exactly what she needed at the moment.”* [P110]

*“When she became a mum but she couldn’t see the bonding, she couldn’t feel it. After the feedback, watching the videos, she was able to see the bond….she could feel the attachment, her bonding with the baby was very strong… she became very confident in seeing how she was interacting with the baby”* [Clinician of P105]

Conversely, because they were very aware of mothers’ anxieties about themselves as parents, some clinicians struggled with the requirement in later sessions to point out one instance per session where the mother had responded less sensitively to her baby. The clinicians spoke about trying very hard to word their feedback on these moments gently and sensitively, with support from their supervisor. They found identifying and feeding back on these moments particularly challenging when mothers were generally responding very sensitively to their baby. Three clinicians expressed that they would prefer to use their clinical judgement to determine mothers for whom flagging points for improvement would not be helpful.

*“Because she lacked so much confidence anyway, I didn’t want to criticise…she is really good and really receptive, and responsive to the baby. I found those a bit uncomfortable to say”.* [Clinician of P111]

##### Sub-theme 2.3 Talking about feelings and emotion regulation

Seven mothers described sharing their anxieties and worries about the filming, the feedback, and their parenting, with the clinician. They described clinicians as responding with reassurance and empathy, normalising their feelings and helping them to let go of some of their anxieties.

*“I did feel that she was supportive and kind of normalised any like things that I was struggling with, or concerned about….like I remember we did a clip like about getting her dressed. She hates getting dressed! And she was just kind of normalising that, and then sort of reassuring me, because at the time I was really funny about her crying.”* [P109]

However, three mothers felt that their clinician could have been more proactive in checking in on how they were feeling throughout the session, and would have liked more space in the sessions to explore more generally how they were feeling about their relationship with the baby. Four mothers mentioned holding back on expressing all of their thoughts and feelings about the filming and feedback to the clinician; conversely, three clinicians mentioned that the mothers they were working with did not express any self-critical thoughts or anxieties and so the clinician had not continued to ask about these in every session.

Nine clinicians described finding the emotion regulation tips a helpful addition to the intervention, particularly the advice on accepting their emotions, taking a break when things felt overwhelming, and connecting with baby’s feelings to help them manage their own. Three mothers also described the tips as helpful; however, six mothers could not recall the clinician sharing tips about emotion regulation with them. It was suggested that written information on emotion regulation could have been a useful supplement. One clinician felt the mother she was working with did not need any tips on emotion regulation and so she did not include them; another two clinicians observed that the additional material on emotion regulation meant that there was a lot to cover in each session. More generally, clinicians described preferring to move away from reading information out from the intervention manual, instead drawing on their clinical experience to put information in their own words and to tailor their conversations to the needs of the specific mother they were working with.

##### Sub-theme 2.4 Who is VIPP-PMH for?

Five clinicians felt that, for future wider delivery within perinatal mental health services, VIPP-PMH would be best targeted at mothers who are lacking confidence in themselves as parents or worried about the bond with the baby, and should be available across the wider age range seen by the service (up to 12 months).

*“I think it would be really helpful directed towards a targeted group…. a mum that’s really struggling with the bonding… think about the flexibility of [baby’s] age.”* [Clinician of P103].

## Discussion

### Feasibility and Acceptability of VIPP-PMH with Young Babies

This was the first time the VIPP intervention has been adapted specifically for babies aged under 6 months. Feedback from both mothers and clinicians indicated that VIPP-PMH was viewed overall as working well with 2 to 6 month old babies. The adapted filming tasks were thought to be developmentally appropriate; however, further thought may need to be given to the task requiring baby to hold a toy on their own. Further, the additional psychoeducation on infant communication was found to be helpful. Although babies’ cues at this age were observed to be subtler than with older babies, with practise and supervisory support, clinicians were able to draw out many moments of connection and communication from the interaction. Mothers described being surprised and gladdened by the richness of their baby’s communication and responsivity to them. Both mothers and clinicians felt the intervention helped mothers to better notice their baby’s cues and respond sensitively. In line with this, the pre-post effect size for observer ratings of mother-infant interaction indicated a medium-sized increase in sensitive responsivity to their child. However, with no control condition this improvement cannot necessarily be attributed to VIPP-PMH. Mother-infant interactional quality has previously been shown to increase between 3 and 6 months old, potentially as infants’ cues become more obvious and mothers have gotten to know their baby better (O’Brien et al. 1989). Nonetheless, the findings align with our feasibility trial in older infants (age 6 to 36 months) whose mothers were experiencing moderate to severe perinatal mental health problems, in which sensitivity increased over time in mothers receiving VIPP-PMH but decreased over time in mothers receiving usual care alone (Barnicot et al. 2022). Further, trials in other populations have shown VIPP to be effective in improving maternal sensitivity (Bakermans-Kranenburg et al. 2003, Juffer et al. 2017).

### Feasibility and Acceptability of VIPP-PMH with Mothers Experiencing Moderate to Severe Perinatal Mental Health Difficulties

This is only the second evaluation of the feasibility and acceptability of VIPP for mothers with moderate to severe or complex perinatal mental health difficulties. As in our previous study (Barnicot et al. 2023), mothers experiencing moderate to severe perinatal mental health difficulties described considerable anxieties about being filmed interacting with their baby, However, with reassurance and support from the clinician, and through receiving positive feedback on the interaction focussed on understanding the baby’s cues and communication, mothers’ anxieties were largely allayed. Further, mothers described feeling more confident in themselves as parents and developing a more positive perspective on their relationship with their child. In line with this, outcome data suggested that mothers’ parenting confidence and their perception of a warm relationship with their child improved from pre- to post-intervention with a large effect size, alongside a small improvement in parenting stress. Most mothers appreciated the opportunity to discuss any difficult feelings with their clinician; however, further thought could be paid to checking in more often with mothers who are known to be particularly anxious. Additionally, whilst clinicians found the emotion regulation tips to be a helpful addition to the intervention in this population, the fact that mothers could largely not remember them, and that clinicians described struggling to fit all the material into the session, suggests that this content requires further consideration. The quantitative data showed a small improvement in maternal emotion regulation from pre- to post-intervention; however it is doubtful that this can be attributed to VIPP. VIPP is an intervention designed primarily for parent-infant interaction, rather than for maternal mental health; previous research has shown that over-emphasis on discussing mothers’ own feelings may detract from intervention benefits for children’s future mental health (Klein-Velderman et at. 2006). Potentially the emotion regulation material could be omitted or presented more briefly, and clinicians could adapt the amount of time spent on this topic according to each mother’s need. Similarly, since clinicians described feedback on instances of less sensitive mother-infant interaction to be potentially unhelpful where a mother was already generally highly sensitive and/or felt judged by this, clinicians could be facilitated to use their clinical judgement in deciding whether to feed back on these less sensitive moments.

### Strengths and Limitations

Following a robust intervention adaptation process, this study evaluated the feasibility and acceptability of VIPP-PMH under ecologically valid conditions: the sample was ethnically diverse, with a range of mental health diagnoses, and was recruited across three different perinatal mental health services. Further, VIPP-PMH was delivered by perinatal mental health clinicians in the context of their everyday job role, as it would be if the intervention is rolled out in the future. The findings are limited by the small sample and lack of control condition, meaning that pre-post change in outcomes cannot necessarily be attributed to VIPP-PMH. Further, the sample were highly educated, which does not represent all women using perinatal mental health services (Howard et al. 2022). It is unclear whether there was any selection bias in the women approached to participate by clinicians or in the women consenting to participate.

### Conclusion and Implications for Further Research

The findings suggest that our adaptation of VIPP-PMH is feasible, acceptable and potentially helpful for mothers experiencing moderate to severe and/or complex perinatal mental health difficulties and their 2 to 6 month old infants. Additionally, it can be delivered within perinatal mental health service contexts, provided staff are supported to have the correct training, supervision and protected time for delivery. The feedback has suggested further minor adaptations to increase acceptability and feasibility. Further work is required to test the efficacy of VIPP-PMH in a definitive randomised controlled trial.

## Data Availability

The data that support the findings of this study are available on request from the corresponding author. The data are not publicly available due to privacy or ethical restrictions.

## Acknowledgements

With enormous thanks to the mothers, children and clinicians who took part, and particular thanks to Catherine Furlonge, Charlotte Akinyemi, Daniela Laterza, Emily Hodgson, Karen Baptiste, Karis Dennison, Kirsty Carmichael, Miriam Ahmad, Rachel McGeehin, Savanna Folie, Sue Winslade, Veronica Matjele, Vicky Kavadi-Roach, and Zoe Wood who delivered the intervention sessions. We wish also to thank Camilla Rosan and Marian Bakermans-Kranenburg for their advice on adapting VIPP for younger babies.

